## Supplemental Material 1: Python code for image analysis for "WHotLAMP: A simple, inexpensive, and sensitive molecular test for the detection of SARS-CoV-2 in saliva"

import os
import numpy as np
import cv2
import matplotlib

# identifies image ROIs given path and number of samples
# inputs
#   path: path to file
#   Nsamples: number of samples in image
# outputs
#   huevals: list of hues within pixels corresponding to each identified ROI
#   selectedcomponents: array corresponding to the image segmented into ROIs
def runimg(path,Nsamples):
    #load image and convert to HSV
    img_bgr = cv2.imread(path)
    img_hsv = cv2.cvtColor(img_bgr, cv2.COLOR_BGR2HSV)
    height,width,_ = img_bgr.shape


    #get H (hue), S (saturation), and V (value) components of image
    h = img_hsv[:,:,0]
    s = img_hsv[:,:,1]
    v = img_hsv[:,:,2]

    #set saturation of pixels that are in the top or bottom 10% of value (lightness) to the average saturation of the remaining pixels, to ignore reflections and shadows
    v_thresh = np.percentile(v.flatten(),90)
    s[v > v_thresh] = np.mean(s[v < v_thresh])
    v_thresh = np.percentile(v.flatten(),10)
    s[v < v_thresh] = np.mean(s[v > v_thresh])
    s = cv2.GaussianBlur(s,(5,5),0)

    #threshold image into foreground and background based on saturation value
    thresh,img_fgbg = cv2.threshold(s,0,255,cv2.THRESH_BINARY+cv2.THRESH_OTSU)
    while(np.mean(img_fgbg)/255 > 0.05): #at most 5% should be foreground
        thresh,img_fgbg = cv2.threshold(s,thresh*1.1,255,cv2.THRESH_BINARY)


    #find the connected components of the image. the largest should be the background and the next should be the samples of interest
    n_components,components,stats,centroids = cv2.connectedComponentsWithStats(img_fgbg)
    leftmostpix_all = stats[:,0] #leftmost pixel of each component
    topmostpix_all = stats[:,1] #topmost pixel of each component
    widths_all = stats[:,2] #widths of each component
    heights_all = stats[:,3] #heights of each component
    areas_all = stats[:,4] #size of each component

    #normalize to [0,1]
    leftmostpix_all = leftmostpix_all/s.shape[1]
    topmostpix_all = topmostpix_all/s.shape[0]
    widths_all = widths_all/s.shape[1]
    heights_all = heights_all/s.shape[0]

    #ignore components whose height or width is more than 3x their width or height, respectively
    maxdistortion = 3
    distortions = widths_all/heights_all
    areas_all[(distortions>maxdistortion)] = 0
    areas_all[(distortions<(1/maxdistortion))] = 0

    #ignore components near borders of the image
    areas_all[leftmostpix_all < 0.025] = 0
    areas_all[(leftmostpix_all+widths_all) > 0.975] = 0
    areas_all[topmostpix_all < 0.025] = 0
    areas_all[(topmostpix_all+heights_all) > 0.975] = 0

    #if > 10% of image height or width, discard (probably an edge/shadow). background will also be discarded by this.
    areas_all[widths_all > 0.1] = 0
    areas_all[heights_all > 0.1] = 0


    #sort by area
    sorted_compinds = np.argsort(areas_all)[::-1]

    comp_inds = sorted_compinds[0:Nsamples] #top Nsamples components

    components[~np.isin(components,comp_inds)] = 0
    selectedcomponents = np.zeros(components.shape,dtype=int)

    huevals = []
    img_component_colors = np.zeros(img_bgr.shape,dtype=np.uint8)
    for si in range(Nsamples):
        sample = (components == comp_inds[si])
        sample_filled = (cv2.floodFill(sample.astype(np.float32),None,(0,0),-1)[1] != -1) #fill interior of sample
        sample = sample_filled
        medhue = np.median(h[sample])
        stdhue = np.std(h[sample])
        selectedcomponents[sample_filled] = si+1
        huevals.append(h[sample])
        img_component_colors[sample_filled,:] = np.median(img_bgr[sample_filled],0)

    return huevals,selectedcomponents
