## Supplementary Material 2: WHotLAMP protocol for "WHotLAMP: A simple, inexpensive, and sensitive molecular test for the detection of SARS-CoV-2 in saliva"

1. In a 1.7 mL microfuge tube with a 2x3 mm piece of Whatman No. 1 paper glued with Kwik-Sil™ to the side, 1 mm from the bottom, mix in:
    - 100 μL saliva
    - 50 μL RNAlater™ (R0901, Sigma) or 'Homebrew' (see below)
    - 25 μL Proteinase K (10 mg/mL; PB0451, BioBasic)
2. Pipette up and down ~20x to mix well
3. Incubate at ~26ºC for 5 min (but not below 22 ºC)
4. Incubate at 95ºC for 5 min
5. Remove liquid by decanting or using a pipette
6. Add 1 mL Wash Buffer and **invert vigorously** 20 times
7. Incubate at room temperature for 1 min
8. Remove liquid by decanting or using a pipette
9. Add 1 mL Wash Buffer, incubate at room temperature for 1 min, then remove liquid (this is a brief rinse) leaving as little Wash Buffer as possible
10. Add 50 μL of LAMP rxn mix using ZI-1 primers. (If a control reaction for RNA extraction from saliva is desired, use RAB7A primers instead)
11. Take photo of tube(s) as baseline
12. Incubate 65ºC for 20 min
13. Cool to room temperature (or to 4ºC to maximize color difference between negative and positive reactions)
14. Examine tubes visually for pink (negative) or yellow (positive) color change

**Wash Buffer**

| | |
|---|---|
| 1 mM Tris-Cl pH 8.0 | T4661-100G (Sigma) |
| 0.1 mM EDTA pH 8.0 | E9884-100G (Sigma) |
| 0.1 % Tween-20 | P9416-50ML (Sigma) |

**LAMP mix (per reaction)**

| | |
|---|---|
| 25 µL | 2x WarmStart® Colorimetric LAMP mix (NEB, M1800L) |
| 5 µL | 400 mM guanidine hydrochloride (G3272, Sigma, made fresh from powder) |
| 5 µL | ZI-1 10x primers (16 µM FIP/BIP, 2 µM F3/B3, 4 µM LF/LB) |
| 15 µL | water |

**Homebrew RNAlater:**

| | |
|---|---|
| 3.54 M ammonium sulfate | A4418-500G (Sigma) |
| 16.6 mM sodium citrate | C8532-500G (Sigma) |
| 13.3 mM EDTA | E9884-100G (Sigma) |

**Primers**

| | |
|---|---|
| ZI-1-F3 | GGATACAACTAGCTACAGAGAA |
| ZI-1-B3 | CCACAAGTTACTTGTACCATAC |
| ZI-1-FIP | TTGGTAAAGAACATCAGAACCTGAGGCTGCTTGTTGTCATCTC |
| ZI-1-BIP | CCACCACAAACCTCTATCACCTAACCCTCAACTTTACCAGAT |
| ZI-1-LF | AAGTCATTGAGAGCCTTTGC |
| ZI-1-LB | GTGGTTTTAGAAAAATGGCATTCCC |

| | |
|---|---|
| RAB7A-F3 | ACAGGCCTGGTGCTACAG |
| RAB7A-B3 | CTGCAGCTTTCTGCCGAG |

| | |
|---|---|
| RAB7A-FIP | CAATCGTCTGGAACGCCTGCTCCCTACTTTGAGACCAGTGC |
| RAB7A-BIP | AAGCAGGAAACGGAGGTGGAGGCCCGGTCATTCTTGTCC |
| RAB7A-LF | ACGTTGATGGCCTCCTTG |
| RAB7A-LB | TGTACAACGAATTTCCTGAACC |

Notes:

- For gluing Whatman No. 1 paper, use as little Kwik-Sil™ as possible to maximize surface area exposed to saliva mixture. Ideally, allow Kwik-Sil to cure overnight before using.
- We found item https://cardholepunch.com/products/custom-loyalty-card-hole-punch?variant=7789589790786 (custom shape ID: ND15) worked very well to rapidly punch out Whatman No. 1 pieces, with the tapered end exposed to Kwik-Sil and facing the top of the tube.
- The volume of saliva can be larger than 100 μL as long as the other components are scaled accordingly.
- We have found that incubation of the saliva mixture with Whatman paper can occur between 26ºC and 37ºC. Our testing was consistent at 26ºC. Lower incubation temperatures (e.g. <22ºC) may lead to unreliable results, presumably because the activity of Proteinase K is too low.
- The washing steps are important to remove residual RNAlater soaked in the Whatman paper which can affect the LAMP reaction.
- Use of freshly prepared guanidine hydrochloride solution in the LAMP reaction increases the sensitivity of the colorimetric assay.
- For WHotLAMP assays in 1.7 mL microfuge tubes, the 20 min incubation time at 65ºC is important. Extended incubation times (e.g. >25 min at 65ºC) can lead to non-specific colorimetric change in the negative control. For saliva samples with high SARS-CoV-2 titers, a colorimetric change can be detected in as little as 10 min at 65ºC.
- Cooling tubes after the 65ºC incubation step to ~4ºC helps to bring out the color contrast between positive and negative colorimetric LAMP results.
